# Linguistic fairness in the U.S.: The case of multilingual public health information about COVID-19

**DOI:** 10.1101/2021.09.27.21264211

**Authors:** Damián E. Blasi, Vishala Mishra, Adolfo M. García, Joseph P. Dexter

**Affiliations:** Data Science Initiative, Harvard University, Cambridge, MA 02138; Department of Human Evolutionary Biology, Harvard University, Cambridge, MA 02138; Max Planck Institute for the Science of Human History, Jena, Germany; Linguistic Convergence Laboratory, HSE University, Moscow, Russian Federation; Multidisciplinary Research Unit, Madras Medical College, Chennai, India; Cognitive Neuroscience Center (CNC), Universidad de San Andrés, Buenos Aires, Argentina; National Scientific and Technical Research Council (CONICET), Buenos Aires, Argentina; Departamento de Lingüística y Literatura, Facultad de Humanidades, Universidad de Santiago de Chile, Santiago, Chile; Global Brain Health Institute, University of California, San Francisco, San Francisco, CA 94143

**Keywords:** COVID-19, health equity, health literacy, linguistic fairness, readability

## Abstract

Lack of high-quality multilingual resources can contribute to disparities in the availability of medical and public health information. The COVID-19 pandemic has required rapid dissemination of essential guidance to diverse audiences and therefore provides an ideal context in which to study linguistic fairness in the U.S. Here we report a cross-sectional study of official non-English information about COVID-19 from the Centers for Disease Control and Prevention, the Food and Drug Administration, and the health departments of all 50 U.S. states. We find that multilingual information is limited in many states, such that almost half of all individuals not proficient in English or Spanish lack access to state-specific COVID-19 guidance in their primary language. Although Spanish-language information is widely available, we show using automated readability formulas that most materials do not follow standard recommendations for clear communication in medicine and public health. In combination, our results provide a snapshot of linguistic unfairness across the U.S. and highlight an urgent need for the creation of plain language, multilingual resources about COVID-19.

---

**I**n culturally and linguistically diverse countries, the availability of medical information in multiple languages is a key determinant of health equity, but efforts to increase linguistic fairness in U.S. healthcare remain suboptimal (1–3). Over the past year, the COVID-19 pandemic has required rapid dissemination of critical public health information across the country. Such communication efforts have been complicated by longstanding gaps in the health literacy of the U.S. population (for instance, only 12% of U.S. adults scored as proficient in the 2003 National Assessment of Health Literacy (4)). Similar issues have been identified for information about COVID-19, and individuals with low health literacy report reduced preparedness for the pandemic (5–9). The situation is even more challenging for non-native English speakers; when combined with low health literacy, limited English proficiency is associated with poor health status in the U.S. (4, 10, 11). Accordingly, public guidance about COVID-19 should be equitable in its language coverage and use accessible communication strategies. Moreover, the extent to which these dual imperatives are being met provides a valuable benchmark for the state of linguistic fairness in the country.

The U.S. population includes 48 million immigrants, a figure that is projected to increase to at least 75 million by 2065 (12). Reflecting the diversity of both its immigrant and indigenous populations, the U.S. is a deeply multilingual country. Of its more than 330 million residents, 67 million speak a language other than English at home (13), and 25 million have limited English proficiency (an increase of 156% since 1980) (14). Aside from English, by far the most widely spoken language is Spanish (40 million speakers), followed by Chinese (3.3 million), Tagalog (1.7 million), Vietnamese (1.5 million), and French (1.2 million) (13). The quality of many health resources for non-English users, however, is far from ideal (15, 16), contributing to health disparities nationwide and amplifying structural inequalities accentuated by the pandemic (17–19).

U.S. residents receive information about COVID-19 from a diverse selection of official sources, ranging from federal agencies such as the Centers for Disease Control and Prevention (CDC) and Food and Drug Administration (FDA) to state, county, and local health departments. Although federal guidance is subject to the requirements of the Plain Language Act and dissemination of actionable, easy-to-use health information is a major goal of the Healthy People 2030 initiative (3), the accessibility of official COVID-19 information has often been limited (20–24). Compounding these issues is a lack of uniform standards for the provision of multilingual resources, as well as the Trump administration’s rollback of a federal rule requiring that patients be informed of their right to language interpretation services (25).

In addition to its immediate relevance for improving the equity of the pandemic response, a thorough understanding of the availability and accessibility of non-Anglophone COVID-19 guidance is likely to have longer-term implications. The scope of material written about COVID-19 is unprecedented, and the existence of extensive guidance from all U.S. states creates an opportunity for comparative evaluation of linguistic fairness. Moreover, the availability and quality of Spanish-language guidance is of particular importance because of the size of the user base and the disparate impact the pandemic has had on the Latinx community (26, 27). To this end, we undertook a two-part analysis, first of the equity of language coverage in state-level guidance about COVID-19 and second of the readability of Spanish information from both federal and state sources. We find that many states have substantial linguistic communities not served by existing guidance and that most Hispanophone information, regardless of source, is not in compliance with well-known recommendations for clear communication.

## Results

### Prevalence of baseline resources

Although automated translation services such as Google Translate have substantial limitations (28), providing a machine translation plug-in is a straightforward step to increase the accessibility of a website. We found, however, that only 30 of 51 (59%) health department websites included an option for automated translation of information about COVID-19 (Table 1). Similarly, just 29 (57%) websites referenced or linked to the CDC’s large collection of multilingual resources written for the general public, and 34 (67%) included at least one video in American Sign Language (Table 1).

**Table 1.** Prevalence of key resources across all states and the District of Columbia. Resources were tabulated from a review of official health department websites.

| Resource or Tool | Number (%) of Websites |
| --- | --- |
| Automated translation | 30 (59%) |
| CDC multilingual resources | 29 (57%) |
| American Sign Language videos | 34 (67%) |

### Linguistic coverage of COVID-19 guidelines across U.S. states

We then considered the extent to which state-level information about COVID-19 serves different linguistic communities. In particular, we sought to determine whether the availability of public health guidelines is demographically fair (i.e., if it can be accounted for primarily by the number of users of a given language). For each language and state, we cross-referenced the number of speakers (as reported in the U.S. Census Bureau’s American Community Survey (13)) with the presence of COVID-19 materials for that community (Materials and Methods). Given that Spanish COVID-19 guidelines are available for almost all states (48 of 51 websites), we focused exclusively on speakers of languages other than English and Spanish. We modeled the availability of COVID-19 guidelines for a language *j* in state *i* following a Bayesian generalized linear model (Materials and Methods):

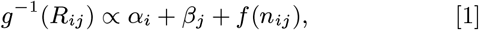

where *R*_*ij*_ is a a binary variable (presence or absence of COVID-19 guidelines), *α* and *β* are group-specific effects for state and language, respectively, *n*_*ij*_ is the corresponding number of speakers, *f*(·) is a smooth function to be learned from the data, and *g*(·) is the logistic function.

Results of the analysis are summarized in Fig. 1. When considering size alone, only large language communities with 50,000 or more individuals are predicted to be covered. Language and state conditional effects also account for a substantial amount of variance, and large states with many small linguistic communities tend to score worse in the fairness of their coverage. Several East African and Southeast Asian languages are comparatively better represented, whereas some Western European languages (German, Dutch, Greek, and Italian) and a handful of large West African languages (Igbo, Yoruba, and Kru) and Navajo are less well-covered, other factors being equal. The users of these languages tend to be proficient in English, which might explain why specific health guidelines were not developed for them. We tested this hypothesis on a subset of the Census data for which the number of users who “speak English less than very well” was indicated (67% of the full dataset). Bayesian model selection corroborated that it is the number of users with limited English proficiency - rather than the total number of users - which better predicts the presence of COVID-19 guidelines (Materials and Methods).

**Fig. 1.**
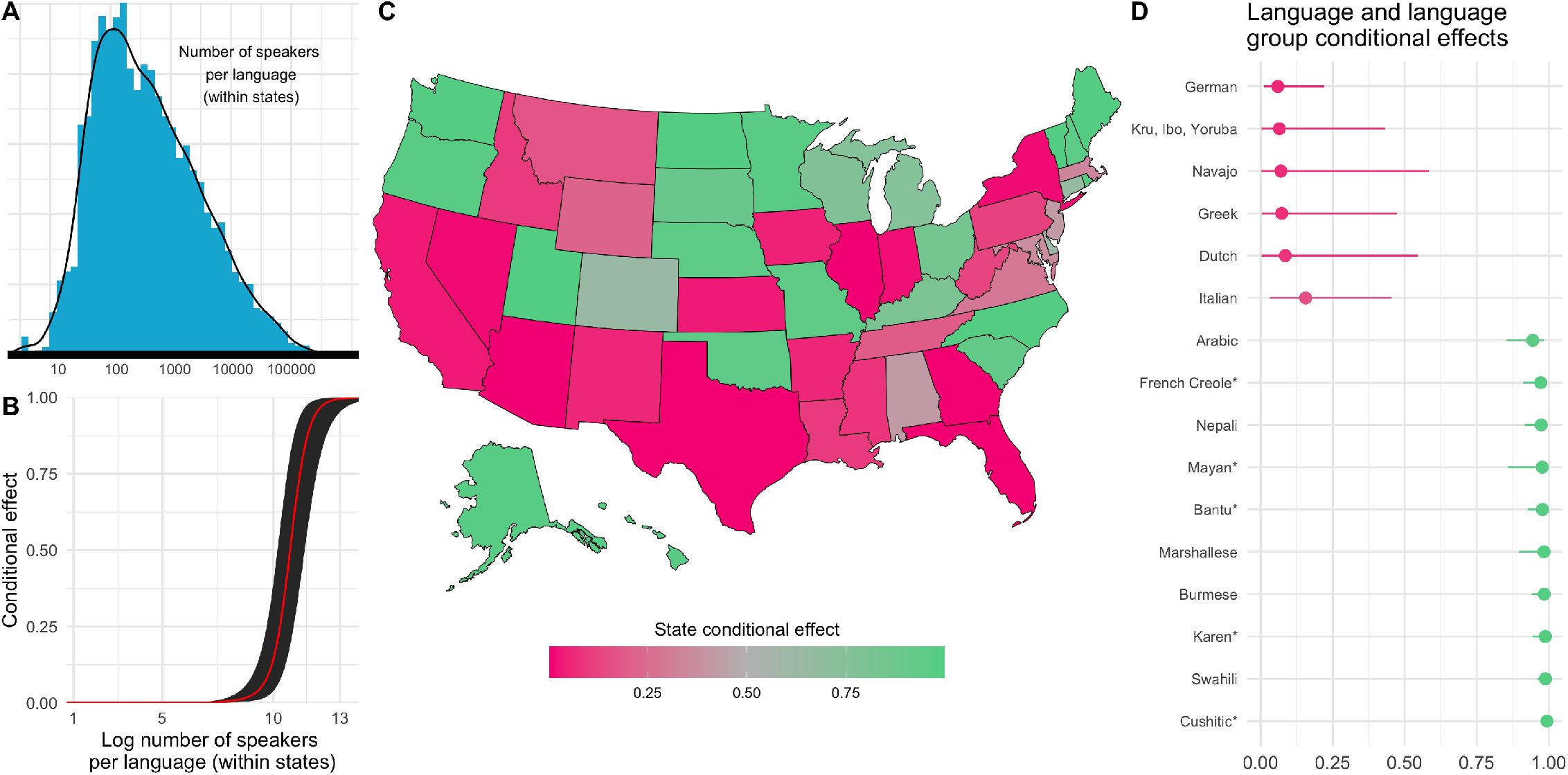
Linguistic diversity and fairness of official COVID-19 information provided by U.S. states. **A** Distribution of number of users per language within states. **B** Conditional effect of (log) number of users of a language on presence of COVID-19 guidelines. **C** U.S. map depicting state-specific effects on presence of COVID-19 guidelines. **D** Bayesian 95% credible intervals for language conditional effects on presence of COVID-19 guidelines.

### Readability of Spanish-language COVID-19 materials

Availability of official guidance in a particular language does not guarantee accessibility and ease of use. Although evaluation of all resources in our multilingual dataset would be challenging, a restricted analysis of the Spanish documents is useful for several reasons. There are more speakers of Spanish in the U.S. than of all other non-English languages combined, and Spanish-language information about COVID-19 is provided by most official sources (Fig. 1). Given the large user base and the wide availability of both manual and automatic English-to-Spanish translation, it is likely that the quality of Spanish resources from the CDC, FDA, and state health departments represents an effective upper bound for the quality of official materials in other languages.

To approximate the text difficulty of Spanish-language COVID-19 information as part of a rapid response to the pandemic, we applied four automated readability formulas to our corpora of public health documents (Materials and Methods). Readability formulas attempt to predict the difficulty of a text, usually expressed as a grade level, and have been used to pinpoint important limitations in Anglophone public health information about COVID-19 (20, 21, 23). These formulas are based on the assumption that, other things being equal, longer linguistic units, such as words and sentences, hinder comprehension. For each of the metrics considered, we found that the vast majority of CDC, FDA, and state documents exceeded an eighth-grade reading level (Fig. 2), which is the CDC, National Institutes of Health, and American Medical Association’s recommended maximum difficulty for health information written for the general public (29–31). Although readability formulas should not be treated as a substitute for reading comprehension data obtained from surveys or focus groups, these results suggest an urgent need for further investigation and development of more accessible communication strategies.

**Fig. 2.**
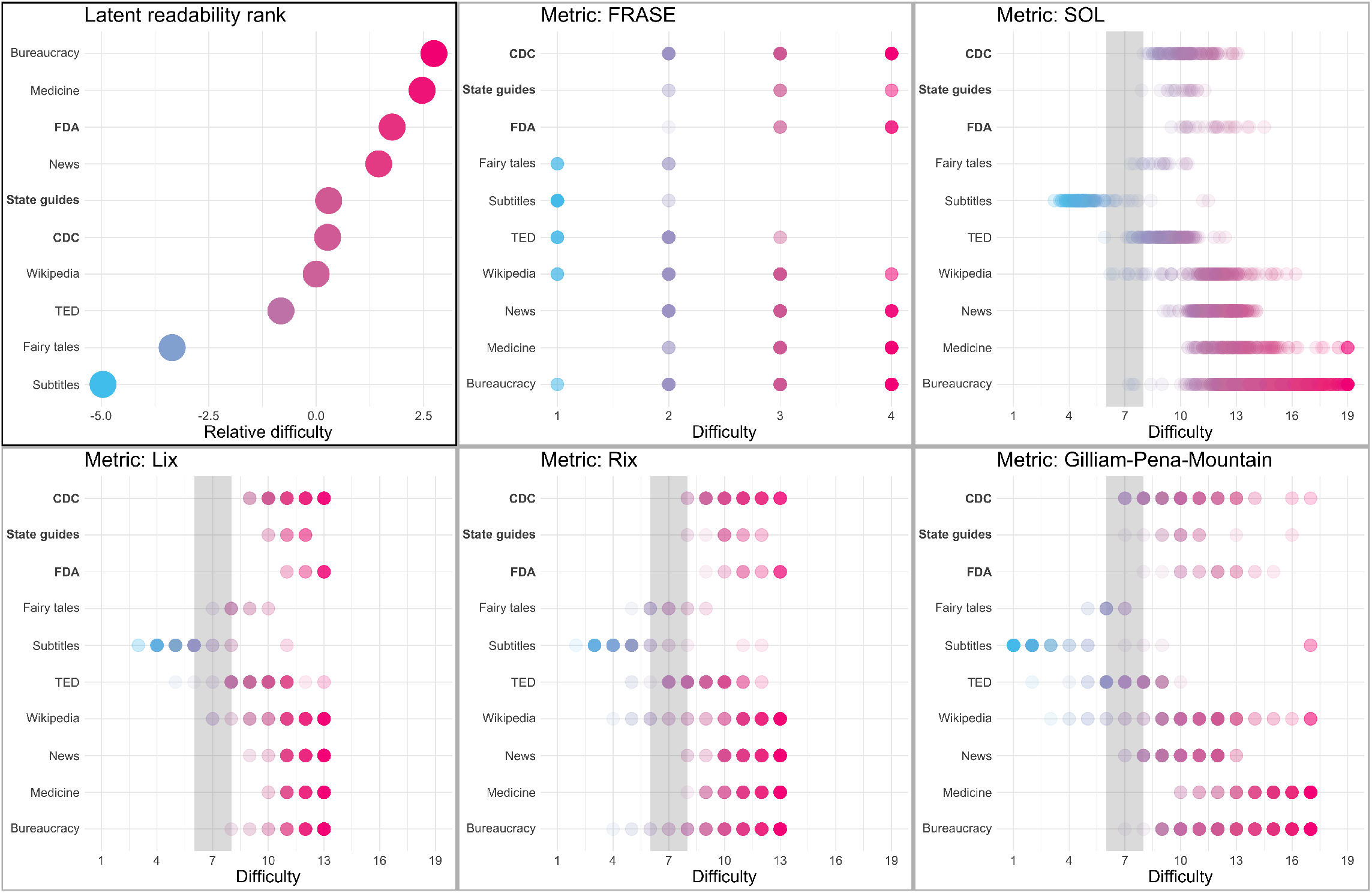
Readability of official Spanish-language information about COVID-19. The figure shows difficulty predictions for the public health and benchmark corpora from five readability metrics - FRASE Graph (FRASE), SOL, Läsbarhetsindex (Lix), Rate Index (Rix), and Gilliam-Pena-Mountain Graph (Gilliam-Pena-Mountain) - as well as a latent rank. The gray bars indicate the recommended reading level for medical information written for the general public (grades 6-8).

Readability measures are not without problems when used as unbiased estimates of overall comprehension and reading level (32–35), in particular for languages other than English (36). Nevertheless, they may provide partial information about the difficulty of one text relative to another, as word length and sentence length - the basis for most readability formulas - are implicated in reading ease and speed (37, 38). To contextualize the results obtained for the public health documents, we also applied the readability metrics to thirteen other corpora covering a wide range of Spanish prose texts of varying genre and difficulty; these corpora include fairy tales, television and film subtitles, transcripts of TED talks, Wikipedia articles, news articles, and medical and bureaucratic documents. We compared the readability of the CDC, FDA, and state materials to the other corpora using all five measures (Fig. 2), and we inferred a latent rank from a Plackett-Luce model (Materials and Methods). As shown in the top left panel in Fig. 2, CDC and state pages were found to be more difficult than the simplest corpora (subtitles, fairy tales, and TED talk transcripts). The FDA documents ranked among the most complex corpora (news, medicine, and bureaucracy).

## Discussion

Here we analyzed the U.S.-wide distribution of COVID-19 guidelines written in languages other than English. The emerging picture is that official COVID-19 information is available for languages with many users who have limited English proficiency but scarce for smaller linguistic communities. While this trend is modulated by the state and the specific language under consideration, the sheer numbers are disheartening (21). Roughly half of all individuals (47%, or 4.2 million people) who are not proficient in either English or Spanish lack access to COVID-19 guidelines in their native language (Materials and Methods). Compounding this inequity in language coverage is the limited availability even of baseline, easy-to-implement resources for non-English users; for instance, more than 40% of state health departments do not offer automated translation of their online information about COVID-19. Although guidance in Spanish is widely available (coverage by the CDC, FDA, and 48 out of 51 state websites), we find that the readability of this material far exceeds the recommended eighth-grade level according to five automated metrics (29–31).

Despite the well-known limitations of readability formulas, including focus on shallow linguistic features and often incomplete validation (32–35), we were able to confirm their reliability for relative assessment of text difficulty; in particular, all metrics considered yielded intuitively reasonable difficulty rankings across a diverse collection of Spanish prose. While our results identify points of urgent concern for the ongoing pandemic response, further work is needed to expand the scope of evaluation to include multimedia resources and traditional, offline media, as well as guidance issued by municipal and county health officials (39). Such an expanded evaluation should seek to integrate statistical and computational evidence with empirical assessments of comprehension and usability testing of multilingual resources (9, 40).

Despite extensive efforts to address the COVID-19 “infodemic” (41), proliferation of misinformation, especially about COVID-19 vaccines, remains a persistent issue (42, 43). Our results thus point to a concerning combination of structural factors - incomplete coverage of multilingual resources, restricted availability of plain language materials, and, as documented previously, poor media ecology - that make it particularly challenging for U.S. residents with limited English proficiency to access trustworthy information. In light of the limitations of official information, many private groups have worked during the pandemic to improve health equity through crowdsourced translation projects, creation of accessible, multilingual public service announcements, and other initiatives (39, 44, 45). These efforts could serve as a blueprint for future efforts that address the challenges identified by our analysis and increase the accessibility of health information for diverse audiences.

## Materials and Methods

### Annotation of multilingual state resources

Between March 1, 2021 and March 15, 2021 we reviewed the websites of the health departments of all 50 states and the District of Columbia for availability of non-English information about COVID-19. For each state we recorded the number of languages represented, as well as the presence or absence of an option for automated translation of the website, reference to the CDC’s large collection of multilingual resources (https://www.n.cdc.gov/pubs/other-languages?Sort=Lang%3A%3Aasc), and American Sign Language videos. To minimize subjectivity in the annotation process we did not assess content or accessibility of resources; a state received credit for a language if its website included any content (whether original or linked directly from an outside resource) in that language.

State-level information about the number of users of each language considered was obtained from the 2009-2013 American Community Survey of the U.S. Census Bureau (https://www.census.gov/data/tables/2013/demo/2009-2013-lang-tables.html). This information was used in our language coverage model and to estimate the total number of individuals not served by state COVID-19 guidance. The linguistic annotation in the Census data is of widely varying quality. While language names often are given directly, a number of coarser language labels are used as well (e.g., by aggregating all users of a language family under a single label or by using coarse regional terms). Whenever possible, we cross-referenced our dataset with the Census information using the widest language label available, so as to overestimate the coverage of state guidelines (see Appendix for details).

### Linguistic coverage model

We implemented the model described in Eq. (1) in a Bayesian framework, utilizing the No-U-Turn Sampler as our Markov Chain Monte Carlo algorithm of choice (46). We used the default weakly informative priors provided by the brms package, version 2.15.0 (47). Posterior predictive checks revealed that the models succeeded in capturing the distribution of the observed data. We then considered the subset of data for which information about English proficiency was available. On this dataset we deployed the model described above, which we contrasted with an equivalent model in which the predictor *n*_*i*_*j* of Eq. (1) was replaced by the number of users of that language with limited English proficiency (plus one, as some languages are reported to have no proficient English users). We performed model selection using the difference in expected log pointwise predictive density between the two models (48), which yielded -15.3 (SE=5.3), favoring the second model over the first one.

### Readability and text complexity analysis

We analyzed the readability and text complexity of online public health information about COVID-19 and 14 benchmark corpora containing a diverse selection of Spanish prose. The public health corpora were drawn from three sources, the CDC, FDA, and state health departments. The CDC corpus contains 153 web pages from the organization’s main Spanish-language website (all pages under the “Su salud” and “Vacunas” tabs of https://espanol.cdc.gov/coronavirus/2019-ncov/index.html, except for landing pages and those with primarily non-textual content). The FDA corpus contains 26 web pages, such as lists of frequently asked questions and vaccine information sheets, from https://www.fda.gov/about-fda/fda-en-espanol/enfermedad-del-coronavirus-covid-19. The state corpus contains 21 web pages from the 12 states that provided a list of frequently asked questions or similar material in Spanish and rank in the top 20 by fraction of Spanish speakers (Texas, California, New Mexico, Florida, Arizona, New Jersey, Illinois, Rhode Island, Utah, Oregon, Washington, and Kansas). CDC and FDA web pages were scraped on March 24, 2021; state web pages were scraped on April 24, 2021. Readability Studio Professional, version 2020 (Oleander Software) was used for text preprocessing, such as removal of headers and footers, figure captions, and other extraneous content.

The benchmark corpora include Spanish translations of 21 of Grimm’s Fairy Tales (https://www.grimmstories.com/es/grimm_cuentos/favorites), as well as 11 sections of the Spanish Unannotated Corpora (SUA) resource (https://github.com/josecannete/spanish-corpora). These corpora are large (the SUA contains approximately 3 billion tokens in total) and cover a range of document types, including Wikipedia and news articles, government documents, and transcripts of speeches. For our analysis we downsampled the corpora by choosing 200 documents at random from each corpus.

We applied five standardized readability formulas, the FRASE Graph, Gilliam-Peña-Mountain Graph, Läsbarhetsindex (Lix), Rate Index (Rix), and SOL, to the public health documents and benchmark corpora. Each formula was developed or adapted specifically for Spanish and has been applied in previous health literacy studies (30). The output of the FRASE formula is a categorical estimate of text difficulty (beginning, intermediate, advanced intermediate, advanced); the output of the other four formulas is a predicted grade level. For our analysis, texts scoring as “13+” by Lix or Rix or “19+” by SOL were reassigned scores of 13 and 19, respectively, and texts scored as “too difficult to be classified” by Gilliam-Peña-Mountain were reassigned a score of 17 (the maximum possible value). Readability scores were calculated using Readability Studio Professional.

Finally, we inferred a latent readability rank for each corpus class across all five readability measures using the R package `PlackettLuce` implementation of the Plackett-Luce model, which represents rankings through a continuous latent variable (49).

## Data Availability

Data is available from the corresponding authors on request.

## ACKNOWLEDGMENTS

We thank Fernando Alva Manchego and Pramit Chaudhuri for helpful discussions. D.E.B. was supported by a Harvard Data Science Fellowship and the HSE University Basic Research Program, funded by the Russian Academic Excellence Project “5-100.” A.M.G. was supported by CONICET; ANID, FONDECYT Regular (grant numbers 1210176 and 1210195); and Programa Interdisciplinario de Investigación Experimental en Comunicación y Cognición (PIIECC), Facultad de Humanidades, USACH. J.P.D. was supported by a Harvard Data Science Fellowship and a CoronaVirus Facts Alliance Grant from the Poynter Institute.

## References

1. EM Feuerherm, RE Showstack, MG Santos, GA Martinez, HE Jacobson, Language as a social determinant of health: Partnerships for health equity in Extending Applied Linguistics for Social Impact: Cross-Disciplinary Collaborations in Diverse Spaces of Public Inquiry, eds. DS Warriner, ER Miller. (Bloomsbury Publishing, New York, New York), pp. 125–148 (2021).

2. J Espinoza, S Derrington, How should clinicians respond to language barriers that exacerbate health inequity? AMA J. Ethics 23, E109–E116 (2021).

3. United States Department of Health and Human Services, Healthy People 2030 (https://health.gov/healthypeople) (2020).

4. M Kutner, E Greenburg, Y Jin, C Paulsen, S White, The health literacy of America’s adults: Results from the 2003 National Assessment of Adult Literacy (https://nces.ed.gov/pubs2006/006483.pdf2) (2006).

5. O. Okan, et al., Coronavirus-related health literacy: A cross-sectional study in adults during the COVID-19 infodemic in Germany. Int. J. Environ. Res. Public Heal. 17, 5503 (2020).

6. E Bilancini, L Boncinelli, V Capraro, T Celadin, R Di Paolo, The effect of norm-based messages on reading and understanding COVID-19 pandemic response governmental rules. J. Behav. Econ. for Policy 4, 45–55 (2020).

7. I Montagni, et al., Acceptance of a Covid-19 vaccine is associated with ability to detect fake news and health literacy. J. Public Heal. 2021, 1–8 (2021).

8. MS Wolf, et al., Awareness, attitudes, and actions related to COVID-19 among adults with chronic conditions at the onset of the US outbreak: a cross-sectional survey. Annals Intern. Medicine 173, 100–109 (2020).

9. V Mishra, JP Dexter, Response of unvaccinated US adults to official information about the pause in use of the Johnson & Johnson-Janssen COVID-19 vaccine. medRxiv 2021 (2021).

10. T Sentell, KL Braun, Low health literacy, limited English proficiency, and health status in Asians, Latinos, and other racial/ethnic groups in California. J. Heal. Commun. 17, 82–99 (2012).

11. LC Diamond, DS Reuland, Communicating with Spanish-speaking patients—Reply. J. Am. Med. Assoc. 301, 2327–2328 (2009).

12. A Budiman, Key findings about U.S. immigrants (https://www.pewresearch.org/fact-tank/2020/08/20/key-findings-about-u-s-immigrants/) (2020).

13. United States Census Bureau, American Community Survey (https://www.census.gov/programs-surveys/acs) (2015).

14. J Batalova, J Zong, Language diversity and English proficiency in the United States. Migr. Inf. Source 2016 (2016).

15. E Jacobs, K Karavolos, P Rathouz, T Ferris, L Powell, Limited English proficiency and breast and cervical cancer screening in a multiethnic population. Am. J. Public Heal. 95, 1410–1416 (2005).

16. C Honeycutt, et al., Assessment of Spanish translation of websites at top-ranked US hospitals. JAMA Netw. Open 4, e2037196 (2021).

17. D Khullar, DA Chokshi, Challenges for immigrant health in the USA—the road to crisis. Lancet 393, 2168–2174 (2019).

18. A A-Oraibi, Migrant health is public health: a call for equitable access to COVID-19 vaccines. Lancet Public Heal. 6, e144 (2021).

19. R Khazanchi, C Evans, J Marcelin, Racism, not race, drives inequality across the COVID-19 continuum. JAMA Netw. Open 3, e2019933 (2020).

20. V Mishra, J Dexter, Comparison of readability of official public health information about COVID-19 on websites of international agencies and the governments of 15 countries. JAMA Netw. Open 3, e2018033 (2020).

21. S Khan, A Asif, AE Jaffery, Language in a time of COVID-19: Literacy bias ethnic minorities face during COVID-19 from online information in the UK. J. Racial Ethn. Heal. Disparities 8, 1242–1248 (2021).

22. CM Chen, Public health messages about COVID-19 prevention in multilingual Taiwan. Multilingua 39, 597–606 (2020).

23. ST Lim, M Kelly, S Johnston, Re: ‘Readability of online patient education material for the novel coronavirus disease (COVID-19): a cross-sectional health literacy study’. Public Heal. 190, 145–146 (2021).

24. J Wernimont, Media infrastructure is and has always been a matter of life and death. Items 2021 (2021).

25. K Keith, HHS strips gender identity, sex stereotyping, language access protections from ACA anti-discrimination rule. Heal. Aff. Blog 2020 (2020).

26. MW Hooper, A. Nápoles, EJ Pérez-Stable, COVID-19 and racial/ethnic disparities. J. Am. Med. Assoc. 323, 2466–2467 (2020).

27. M Jacobson, TY Chang, M Shah, R Pramanik, SB Shah, Racial and ethnic disparities in SARS-CoV-2 testing and COVID-19 outcomes in Medicaid managed care cohort. Am. J. Prev. Medicine 000, 1–8 (2021).

28. X Chen, S Acosta, AE Barry, Evaluating the accuracy of Google translate for diabetes education material. JMIR Diabetes 1, e5848 (2016).

29. S Badarudeen, S Sabharwal, Assessing readability of patient education materials: current role in orthopaedics. Clin. Orthop. Relat. Res. 468, 2572–2580 (2010).

30. SA Novin, EH Huh, MG Bange, FK Hui, PH Yi, Readability of Spanish-language patient education materials from RadiologyInfo.org. J. Am. Coll. Radiol. 16, 1108–1113 (2019).

31. Centers for Disease Control and Prevention, Simply Put: A guide for creating easy-to-understand materials (https://www.cdc.gov/healthliteracy/pdf/simply_put.pdf) (2009).

32. J Redish, Readability formulas have even more limitations than Klare discusses. ACM J. Comput. Documentation 24, 132–137 (2000).

33. E Pitler, A Nenkova, Revisiting readability: A unified framework for predicting text quality in Proceedings of the Conference on Empirical Methods in Natural Language Processing, EMNLP ‘08. (Association for Computational Linguistics, USA), p. 186–195 (2008).

34. M Zamanian, P Heydari, Readability of texts: State of the art. Theory & Pract. Lang. Stud. 2 (2012).

35. SA Crossley, S Skalicky, M Dascalu, Moving beyond classic readability formulas: new methods and new models. J. Res. Read. 42, 541–561 (2019).

36. T François, C Fairon, An “AI readability” formula for French as a foreign language in Proceedings of the 2012 Joint Conference on Empirical Methods in Natural Language Processing and Computational Natural Language Learning, EMNLP-CoNLL ‘12. (Association for Computational Linguistics, USA), p. 466–477 (2012).

37. K Rayner, TJ Slattery, D Drieghe, SP Liversedge, Eye movements and word skipping during reading: effects of word length and predictability. J. Exp. Psychol. Hum. Percept. Perform. 37, 514 (2011).

38. BB Kadayat, E Eika, Impact of sentence length on the readability of web for screen reader users in International Conference on Human-Computer Interaction. (Springer), pp. 261–271 (2020).

39. I Feinberg, Building a culture of health literacy during COVID-19. New Horizons Adult Educ. Hum. Resour. Dev. 33, 60–64 (2021).

40. J Howe, C Young, C Parau, G Trafton, R Ratwani, Accessibility and usability of state health department COVID-19 vaccine websites: A qualitative study. JAMA Netw. Open 4, e2114861 (2021).

41. J Zarocostas, How to fight an infodemic. Lancet 385, P676 (2020).

42. G Pennycook, J McPhetres, Y Zhang, J Lu, D Rand, Fighting COVID-19 misinformation on social media: Experimental evidence for a scalable accuracy-nudge intervention. Psychol. Sci. 31, 770–780 (2020).

43. D Scales, J Gorman, K Jamieson, The Covid-19 infodemic—applying the epidemiologic model to counter misinformation. New Engl. J. Medicine 385, 678–681 (2021).

44. The Community Health Literacy Project (https://communityhealthliteracyproject.org/) (2020).

45. Coronavirus Adult Literacy Resources (https://education.gsu.edu/research-outreach/alrc/adultliteracy-coronavirus-resource-links/) (2020).

46. MD Hoffman, A Gelman, The No-U-Turn sampler: adaptively setting path lengths in Hamiltonian Monte Carlo. J. Mach. Learn. Res. 15, 1593–1623 (2014).

47. PC Bürkner, brms: An R package for Bayesian multilevel models using Stan. J. Stat. Softw. 80, 1–28 (2017).

48. J Piironen, A Vehtari, Comparison of Bayesian predictive methods for model selection. Stat. Comput. 27, 711–735 (2017).

49. HL Turner, J van Etten, D Firth, I Kosmidis, Modelling rankings in R: the PlackettLuce package. Comput. Stat. 35, 1027–1057 (2020).

