## Appendix for "Linguistic fairness in the U.S.: The case of multilingual public health information about COVID-19"

PNAS
www.pnas.org

### Supplementary Appendix for

#### Linguistic fairness in the U.S.: The case of multilingual public health information about COVID-19

**Damián E. Blasi, Vishala Mishra, Adolfo M. García, and Joseph P. Dexter**

**Corresponding authors: Damián E. Blasi, Vishala Mishra, Joseph P. Dexter**
****

**This PDF file includes:**

Table S1

| Label | Languages | Description |
|---|---|---|
| Guajarati | Gujurati \| Gujarati | Spelling normalization |
| Samoan | Somoan \| Samoan | Spelling normalization |
| Laotian | Lao \| Laotian | Spelling normalization |
| Punjabi | Punjabi \| Panjabi | Spelling normalization |
| Khmer | Khmer \| Mon-Khmer, Cambodian | Spelling normalization |
| Punjabi | Punjabi \| Panjabi | Spelling normalization |
| Purepecha | Purepecha \| Tarascan | Alternative names |
| Serbo-Croatian* | Serbian \| Croatian \| Bosnian \| Serbocroatian | Generalization |
| Mayan* | K'iche \| Q'anjobal \| Mam \| Mayan \| Chuj \| Akateko \| Mayan Languages | Generalization |
| Cushitic* | Cushite \| Somali \| Oromo \| Maay Maay | Generalization |
| Chamic* | Cham \| Rhade \| Jarai | Generalization |
| Persian* | Farsi \| Persian \| Dari | Generalization |
| Chinese* | Chinese \| Mandarin \| Cantonese | Generalization |
| Micronesian* | Micronesian \| Pohnpeian \| Kosraean \| Chuukese | Generalization |
| Bantu* | Kirundi \| Lingala \| Bantu \| Kinyarwanda \| Kinyamulenge | Generalization |
| Karen* | Karen \| Kayah | Generalization |
| Tibetan* | Tibetan \| Bhantanese | Generalization |
| Oto-Manguean* | Mixteco \| Zapoteco del Istmo \| Oto-Manguen | Generalization |
| Sudanic* | Dinka \| Nuer \| Sudanic | Generalization |
| French Creole* | Haitian Creole \| French Creole | Generalization |

**Table S1. Language and language group labels used in the analysis.** "Label" is the summary term for the related languages, language names, and language varieties we aggregated, which are listed under "Languages". The "Description" column summarizes the nature of the normalization process. "Spelling normalization" and "Alternative names" refer to standardization of orthography and nomenclature, respectively; "Generalization" refers to cases in which it was necessary due to incomplete information to identify the most proximate common linguistic denominator across databases.
